# Rapid and scalable preclinical evaluation of personalized antisense oligonucleotide therapeutics using organoids derived from rare disease patients

**DOI:** 10.1101/2023.03.28.23287871

**Authors:** John C. Means, Daniel A. Louiselle, Emily G. Farrow, Tomi Pastinen, Scott T. Younger

## Abstract

Personalized antisense oligonucleotides (ASOs) have achieved positive results in the treatment of rare genetic disease. As clinical sequencing technologies continue to advance, the ability to identify rare disease patients harboring pathogenic genetic variants amenable to this therapeutic strategy will likely improve. Here, we describe a scalable platform for generating patient-derived cellular models and demonstrate that these personalized models can be used for preclinical evaluation of patient-specific ASOs. We establish robust protocols for delivery of ASOs to patient-derived organoid models and confirm reversal of disease-associated phenotypes in cardiac organoids derived from a Duchenne muscular dystrophy (DMD) patient harboring a structural deletion in the dystrophin gene amenable to treatment with existing ASO therapeutics. Furthermore, we design novel patient-specific ASOs for two additional DMD patients (siblings) harboring a deep intronic variant in the dystrophin gene that gives rise to a novel splice acceptor site, incorporation of a cryptic exon, and premature transcript termination. We show that treatment of patient-derived cardiac organoids with patient-specific ASOs results in restoration of DMD expression and reversal of disease-associated phenotypes. The approach outlined here provides the foundation for an expedited path towards the design and preclinical evaluation of personalized ASO therapeutics for a broad range of rare diseases.

## Main

Antisense oligonucleotides (ASOs) are short synthetic nucleic acid molecules that, when designed to be complementary to an intracellular mRNA target, can influence RNA processing and/or impact protein expression levels. In the four decades since their initial application in the laboratory, more than a dozen different antisense therapies have been approved in the United States for the treatment of a variety of diseases^1^. The inherent flexibility of a sequence-based targeting modality, in combination with well-established pharmacokinetic and pharmacodynamic properties, make ASOs a particularly attractive strategy for the treatment of genetic disorders. Moreover, the ability to customize ASO designs to individual patients has major implications for personalized therapeutics. For example, personalized ASOs designed against a splice-disrupting transposon insertion within an intron of the MFSD8 gene in a Batten’s disease patient were recently shown to significantly reduce the frequency and duration of disease-associated seizures^2^. While encouraging, the overwhelming time and cost requirements for the design and preclinical evaluation of personalized ASOs currently precludes this strategy from more widespread adoption.

Induced pluripotent stem cells (iPSCs) have become powerful tools for modeling developmental processes and dissecting disease pathology^3,4^. Recent advancements in the generation of three-dimensional organoids from patient-derived iPSCs have further expanded the potential of these cellular systems^5^. However, current approaches for establishing patient-derived iPSC models can be time consuming and costly. These limitations often prevent patient-derived models from reaching their full therapeutic potential with respect to the patients from which they were generated.

We address several of these challenges through the development of a rapid, robust, and scalable platform for the generation of patient-derived cell models. We describe an iPSC reprogramming pipeline that utilizes cryopreserved peripheral blood mononuclear cells (PBMCs), commonly available from prior genetic testing, and requires only 3 weeks to establish iPSC lines. In addition, we demonstrate that 3-dimensional organoid models generated from patient-derived iPSCs recapitulate disease-associated phenotypes that can be reversed using patient-specific ASOs. The approach we outline, from procurement of patient PBMCs to empirical analysis of ASO effects on organoid function, requires no specialized equipment and less than 8 weeks of hands-on time. We anticipate that implementation of the platform we describe will lead to the rapid development of preclinical ASO leads for the treatment of many rare genetic diseases.

### Generation of patient-derived iPSC lines at scale

Our research institute recently launched a rare disease initiative, “Genomic Answers for Kids”, which aims to sequence the genomes of 30,000 children impacted by genetic conditions over the course of 7 years^6^. Upon enrollment, patients provide a standard blood draw for genome sequencing and residual PBMCs are cryopreserved for future research use. We reasoned that these patient-specific PBMCs could be a valuable resource for building a biobank of patient-derived cellular models of rare genetic diseases. Realizing this potential, however, requires several technical hurdles to be addressed.

Generating iPSCs from patient cells can be achieved through several different techniques. Viral-based approaches (e.g. Sendai virus) can be highly efficient, but the current reagent expense is not amenable to scalability^7^. Episomal plasmid-based approaches are more cost-effective but are less efficient and require more source material^8^. To overcome these limitations, we performed an extensive optimization of multiple iPSC reprogramming strategies, ultimately coalescing these methods into a single basic protocol. Key features of our protocol include a customized reprogramming efficiency cocktail (MEK inhibitor CHIR99021, TGF-*β*/Activin/Nodal receptor inhibitor SB431542, and ROCK inhibitor Y-27632) and a centrifugation step to facilitate attachment to Matrigel-coated plates (See Supplemental Methods). Using this protocol, we are routinely able to reprogram PBMCs into iPSCs, expand iPSC lines, and cryopreserve patient-derived iPSC lines into our biobank within 2-3 weeks (Figure 1a). Moreover, reprogramming reactions are performed in a single well of a 12-well dish which conveniently enables 12 independent reprogramming experiments to be performed in parallel (Figure 1a).

**Figure 1.**
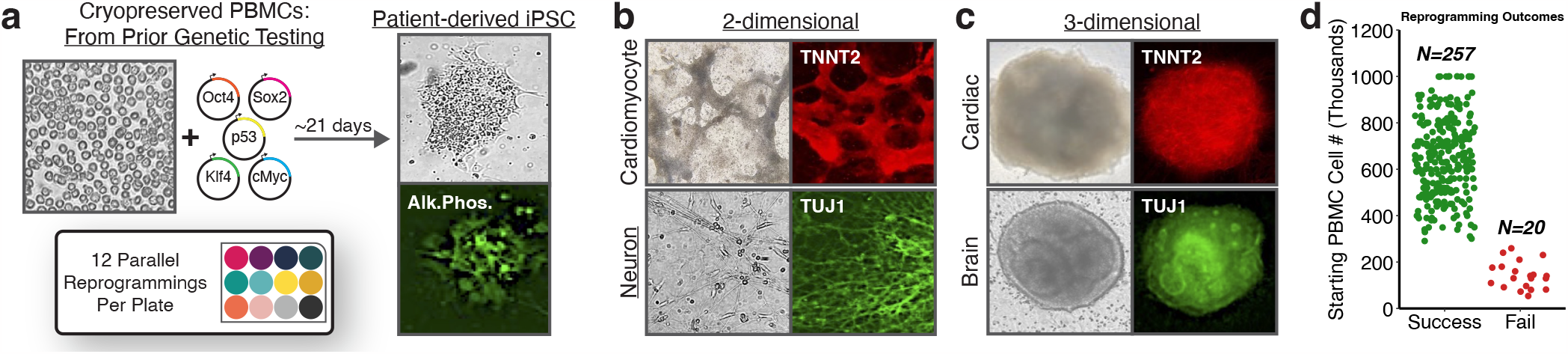
A rapid and scalable platform for the generation of patient-derived cellular models. **a)** Schematic of iPSC reprogramming workflow. **b)** Differentiation of patient-derived iPSCs into 2-dimensional cardiomyocytes and neurons. **c)** Differentiation of patient-derived iPSCs into 3-dimensional cardiac and brain organoids. **d)** Reprogramming outcomes relative to PBMC input cell counts. **b**,**c** representative bright field and fluorescence images from independent differentiation experiments.

To ensure that patient-derived iPSC lines are pluripotent we have evaluated their differentiation potential for a variety of lineages. With an initial focus on 2-dimensional differentiation systems, we found that patient-derived iPSCs could be readily differentiated into both cardiomyocytes and neurons (Figure 1b). We next confirmed the ability of patient-derived iPSCs to generate 3-dimensional organoid models. Again, we found that patient-derived iPSCs could be readily differentiated into both cardiac organoids and whole brain organoids (Figure 1c).

Given the high efficiency and inherent scalability of our reprogramming pipeline, we sought to determine the reliability and robustness of our approach through a campaign of iPSC reprogramming experiments. Over the course of six months we were able to generate >250 patient-derived iPSC lines with >92% success rate (Figure 1d). Importantly, we found that reprogramming outcomes were influenced predominantly by the number of available PBMCs and not by any intrinsic features of a given patient or disease phenotype. More specifically, we were able to generate iPSCs for 100% of patients for which at least 300K PBMCs were available (Figure 1d). Notably, our protocol was also capable of deriving iPSCs from patient fibroblasts (Figure S1). The iPSC reprogramming protocol we outline does not require specialized equipment or high-cost reagents and can be easily implemented in any standard research laboratory.

### Robust genetic perturbation in patient-derived cell models using ASOs

Having harnessed the ability to rapidly generate patient-derived cellular models, we next focused our efforts on performing genetic perturbations in these systems. We focused specifically on ASO technology due to their established use as a laboratory reagent and their recent application towards personalized therapeutics^2^. To optimize our ASO delivery strategy we selected a target gene for which: a) protein expression could be easily quantified, and b) inhibition of gene expression would have a visible impact on cellular phenotypes. Cardiac troponin T (TNNT2) is highly expressed in cardiomyocytes and is required for cardiomyocyte contraction^9–11^. We reasoned that inhibition of cardiac troponin T in iPSC-derived cardiac models would provide an ideal system for evaluating ASO delivery and efficacy.

To explore a diversity of genetic manipulations we designed three distinct ASOs targeting different regions of the cardiac troponin T transcript (Figure 2a). We first designed an ASO targeting the AUG translation start site, anticipating that ASO binding would sterically interfere with ribosome assembly and prevent protein production. Several existing ASO therapeutics function through the modulation of RNA splicing as opposed to direct inhibition of protein expression^12–15^. To ensure that our ASO strategy was capable of influencing RNA splicing we designed two additional ASOs, one targeting a splice donor site and one targeting a splice acceptor site. We synthesized all ASOs, including a negative control ASO with no complementarity to the human genome, as fully modified 2’-O-methyl RNAs.

**Figure 2.**
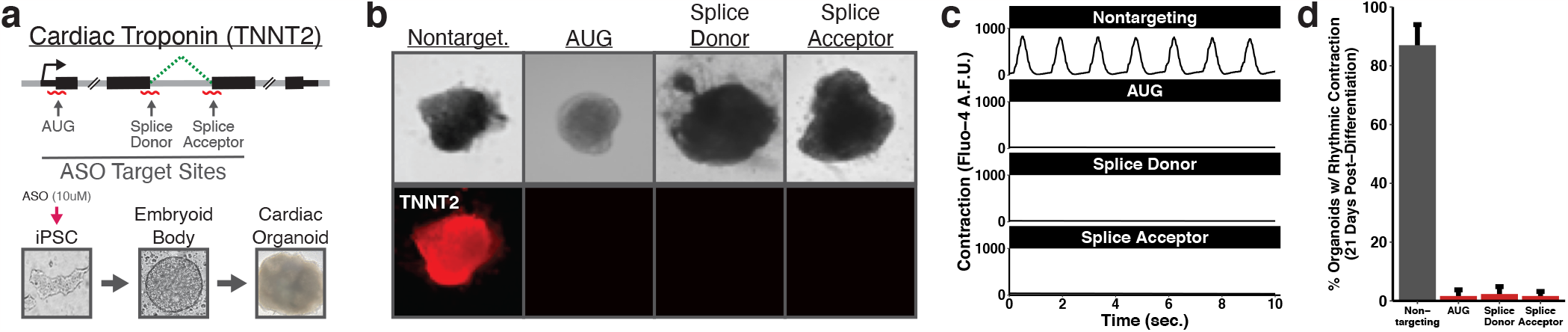
Robust genetic perturbation in iPSC systems using ASOs. **a)** Schematic of design and delivery strategy for ASOs targeting cardiac troponin T. **b)** Cardiac troponin T expression in cardiac organoids derived from ASO-treated iPSCs. **c)** Contraction of individual cardiac organoids derived from ASO-treated iPSCs as determined by intracellular calcium levels. **d)** Summarized contraction of cardiac organoids derived from ASO-treated iPSCs. Error bars in **d** show average and s.e.m. for *n* = 3 counts of 100 independent organoids.

We transfected ASOs into commercially available iPSCs and subsequently initiated differentiation into cardiac organoids (Figure 2a). Cardiac organoids generated from iPSCs treated with the negative control ASO expressed high levels of cardiac troponin (Figure 2b). In contrast, cardiac troponin expression was undetectable in cardiac organoids generated from iPSCs treated with ASOs targeting the AUG translation start site (Figure 2b). Furthermore, cardiac troponin expression was absent in cardiac organoids generated from iPSCs treated with ASOs targeting splice donor/acceptor sites, suggesting that RNA splicing had been altered (Figure 2b).

To quantify the impact of cardiac troponin T inhibition on cardiac organoid function we evaluated calcium fluctuations using the Fluo4-AM calcium indicator dye. Cardiac organoids generated from iPSCs treated with the negative control ASO displayed robust and rhythmic cycles of calcium uptake (Figure 2c, Video S1). However, cardiac organoids generated from iPSCs treated with cardiac troponin T-targeting ASOs displayed extremely weak and arrhythmic levels of calcium uptake (Figure 2c, Videos S2-S4). Cardiac organoid phenotypes remained consistent for at least 21 days following the initiation of cardiac differentiation (Figure 2d). Altogether, these data demonstrate that ASOs can achieve robust genetic perturbation in an iPSC-derived organoid model.

### Evaluating existing ASO therapeutics in patient-derived organoids

Having established methods for charactering ASOs in iPSC-derived organoid models we sought to determine if these models could be suitable for evaluating the efficacy of ASO therapeutics. Among the iPSC lines we generated was one from a Duchenne muscular dystrophy (DMD) patient (Patient 1) harboring a structural deletion of exons 46-53 of the dystrophin gene, resulting in a frameshift in the dystrophin transcript and a loss of dystrophin protein expression (Figure 3a). This deletion is amenable to treatment with an FDA-approved ASO therapeutic designed to skip splicing to exon 45, leading to a dystrophin transcript that lacks genetic information but with a restored reading frame^15^. Although DMD is often described in the context of skeletal muscle, most patients ultimately succumb to cardiac failure making cardiac organoids a highly relevant system for evaluating therapeutic approaches^16,17^. We designed a 2’-O-methyl RNA matching the sequence of the FDA-approved therapeutic to profile its activity in a patient-derived organoid model.

**Figure 3.**
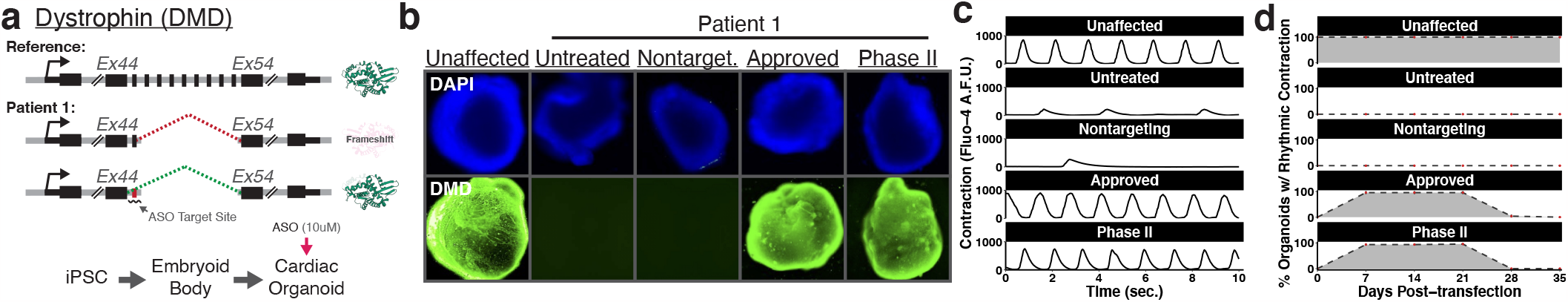
Profiling activity of existing ASO therapeutics in a patient-derived organoid model of disease. **a)** Schematic of design and delivery strategy to profile existing ASO therapeutics for the treatment of DMD. **b)** Dystrophin expression in DMD patient-derived cardiac organoids (patient 1) treated with ASOs matching the sequence of existing ASO therapeutics. **c)** Contraction of individual DMD patient-derived cardiac organoids (patient 1) treated with ASOs matching the sequence of existing ASO therapeutics. **d)** Time course analysis of restored contraction in DMD patient-derived cardiac organoids (patient 1) treated with ASOs matching the sequence of existing ASO therapeutics.

In contrast to our previous experiments in which ASOs were delivered into iPSCs, we reasoned that delivery to differentiated organoids would more closely reflect the ability of the ASOs to function as therapeutics (Figure 3a). We generated cardiac organoids from unaffected iPSCs as well as from iPSCs derived from Patient 1. We validated the resulting cardiac organoids using cardiac troponin expression (Figure S2). In contrast to unaffected cardiac organoids which express high levels of dystrophin, expression was completely absent in organoids generated from Patient 1 (Figure 3b). However, treatment with an ASO matching the sequence of the approved therapeutic restored dystrophin expression to levels comparable to a healthy cardiac organoid (Figure 3b). We also profiled the activity of a second exon 45 skipping ASO that targets a different sequence in the dystrophin transcript and is currently in Phase II clinical trials^18^. Treatment with a 2’-O-methyl RNA matching the sequence of the therapeutic in clinical trials also restored dystrophin expression to levels comparable to a healthy cardiac organoid (Figure 3b).

We next profiled the function of cardiac organoids generated from Patient 1 using the Fluo4-AM calcium indicator dye. Organoids generated from Patient 1 displayed weak and arrhythmic patterns of calcium uptake (Figure 3c, Figure S3, Videos S6-S7). Treatment of patient-derived cardiac organoids with exon 45 skipping ASOs restored the rhythmic calcium fluctuations we observe in healthy cardiac organoids (Figure 3c, Videos S5, S8-9). Moreover, organoid contractions were robust up to 21 days post-transfection (Figure 3d). Organoid contraction was correlated with restored dystrophin expression which also persisted up to 21 days post-transfection (Figure S4). These results indicate that patient-derived organoid models can be used to evaluate the efficacy of ASO therapeutics.

### Preclinical evaluation of personalized ASO therapeutics

The ability to tailor the sequence of an ASO to an individual patient is a powerful feature of this therapeutic modality. We next set out to determine if patient-derived organoid models could be used for the preclinical characterization of novel, patient-specific ASO therapeutics. Included in the cohort of iPSC lines we generated were iPSCs from two additional DMD patients, siblings (Patient 2a and Patient 2b), that inherited a deep intronic mutation in the dystrophin gene that gives rise to a novel splice acceptor site, incorporation of a cryptic exon, and premature transcript termination^19^. We reasoned that this intronic variant would be an ideal candidate target for an ASO therapeutic and designed two ASOs, both 2’-O-methyl RNAs complementary to distinct sequences overlapping the pathogenic variant within the DMD intron (Figure 4a).

**Figure 4.**
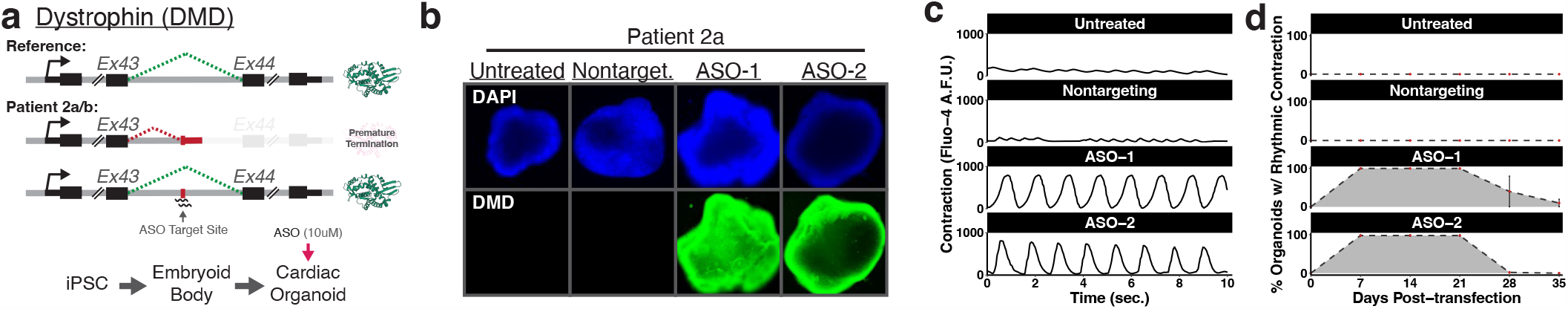
Design and preclinical evaluation of patient-specific ASOs in patient-derived organoids. **a)** Schematic of design and delivery strategy for preclinical evaluation of novel patient-specific ASOs. **b)** Dystrophin expression in DMD patient-derived cardiac organoids (patient 2a) treated with patient-specific ASOs. **c)** Contraction of individual DMD patient-derived cardiac organoids (patient 2a) treated with patient-specific ASOs. **d)** Time course analysis of restored contraction in DMD patient-derived cardiac organoids (patient 2a) treated with patient-specific ASOs.

To evaluate the efficacy of the personalized ASOs we generated cardiac organoids from iPSCs derived from Patient 2a. We validated the resulting cardiac organoids using cardiac troponin expression (Figure S5). As with Patient 1, organoids generated from Patient 2a lacked detectable dystrophin protein expression (Figure 4b). However, treatment with either of the personalized ASOs we designed restored dystrophin protein expression to levels comparable to a healthy cardiac organoid (Figure 4b). Previous reports have shown that restoration of dystrophin expression at levels equivalent to just 20% of healthy individuals has been sufficient to prevent DMD symptoms, suggesting that our ASOs are promising preclinical candidates^20,21^. Notably, cardiac organoids treated with personalized ASOs displayed rhythmic calcium fluctuations identical to those of a healthy cardiac organoid (Figure 4c, Figure S6, Videos S10-S13). As we observed in previous experiments, organoid contractions and dystrophin expression remained robust up to 21 days post-transfection (Figure 3d, Figure S7). Importantly, we obtained indistinguishable experimental results when evaluating the efficacy of the personalized ASOs in cardiac organoids generated from Patient 2b (Figures S5-S8, Videos S14-S17). In summary, these data establish patient-derived organoid models as powerful systems for the design and characterization of personalized ASO therapeutics.

## Discussion

The ability of ASO therapeutics to be customized to individual patients makes their use an appealing strategy for personalized medicine. The recent demonstration that patient-specific ASOs can significantly improve disease-associated phenotypes further supports the application of this technology towards the treatment of rare diseases caused by genetic factors amenable to an ASO-based approach^2^. However, for the full potential of personalized ASOs to be realized an expedited process for the preclinical characterization of candidate ASOs is required. We have developed a rapid, robust, and scalable platform for generating patient-derived cellular models and have validated the utility of these models in the evaluation of patient-specific ASOs. Although significant regulatory considerations remain with regards to the approval of personalized ASOs for use in the clinic, the platform we describe will facilitate widespread preclinical development of personalized ASOs. Increases in the number of prospective patient-specific ASOs will encourage a modernized assessment of the approval process for this unique class of therapeutics.

Patient-derived organoid systems provide a powerful alternative to the use of animal models in the process of drug development. Preclinical studies involving animal models require an average of 6-8 years to complete^21,22^. However, the lifespan for 30% of children born with a rare disease is less than 5 years^23^. As illustrated in this study, patient-derived organoid models recapitulate disease phenotypes and can be generated in less than two months. Although this study describes the use of patient-derived organoids in the evaluation of personalized ASOs, these systems can be adapted for a broad range of drug development approaches (e.g. high-throughput compound screening). Moreover, organoids mimic many key features of human development and offer model systems for exploring diverse aspects of disease biology.

In conclusion, the data described here demonstrate reduction to practice for the use of patient-derived organoid models in the evaluation of personalized therapeutics. The methods and protocols generated in this study are accessible and can be implemented in any standard research laboratory without the need for specialized equipment or high-cost reagents. The widespread ability to generate patient-derived cellular systems will have a significant impact on the understanding of disease mechanisms as well as potential therapeutic avenues for the treatment of many rare diseases.

## Data Availability

All data produced in the present work are contained in the manuscript

## Supplementary Information

### Methods

#### Patient consent

Patients profiled in this study are participants in the Genomic Answers for Kids (GA4K) program at the Children’s Mercy Research Institute. Informed written consent was obtained upon enrollment into the GA4K program.

#### Generation of patient-derived iPSC lines

Control human iPSC line (WTC11 background) was obtained from Coriell Institute (GM25256) (Table S1). Patient iPSCs were generated from peripheral blood mononuclear cells (PBMCs). PBMCs were cultured in StemSpan SFEM II (StemCell Tech, 9605) supplemented with StemSpan erythroid expansion supplement (StemCell Tech, 2692) and 1X Antibiotic-Antimycotic (Gibco, 15240062) for 2-5 days prior to reprogramming using episomal plasmids. Episomal plasmids expressing Oct3/4, Sox2, Klf4, L-Myc, Lin28, P53, and EBNA1 (pCE-mP53DD (41856), pCXB-EBNA1 (41857), pCE-HUL (41855), pCE-hsk (41814), pCE-hOCT3/4 (41813) (Addgene) were introduced into PBMCs by electroporation using the Lonza 4D-Nucleofecter^™^. The nucleofected PBMCs were transferred to a Matrigel (Corning, 354277) coated plate and maintained in StemSpan SFEM II plus 10*μ*M Rock inhibitor. Two days after nucleofection stem cell media 1 (SCM1) composed of ReproTeSR (StemCell Tech, 5926), 10*μ*M Rock inhibitor (Y-27632 dihydrochloride) (Tocris, 1254), 0.5*μ*M PD0325901 (StemCell Tech, 72184), 6*μ*M CHIR99021 (StemCell Tech, 72054), 2*μ*M SB431542 (StemCell Tech, 72234), 250mM sodium butyrate (StemCell Tech, 72242), 50*μ*g/mL ascorbic acid (Sigma, A8960), 10ng/mL bFGF (StemCell Tech, 78003), and 1X Antibiotic-Antimycotic was added to each well without removing any media and centrifuged at 50xg for 30 min and placed back in the incubator for 2 days. After 2 days media was removed and fresh SCM1 added. The next day SCM1 was added without removing media and the following day all media was removed and fresh SCM1 added. This was repeated for 1 week. After week 1 media was changed to SCM2 composed of SCM1 minus SB431542 and media was changed exactly as described for SCM1. iPSC colonies appeared 5-7 days after nucleofection. Once iPSC colonies appeared they were allowed to grow in mTeSR1 media supplemented with a cocktail mix of 400nM sodium butyrate and 100*μ*g/mL ascorbic acid. Once colonies were established ReLeSR (StemCell Tech, 5872) was used to select colonies. The standard timeline for reprogramming and expansion was 2-3 weeks under optimal conditions. A summary of iPSC lines generated from DMD patients is shown in Table S1.

#### ASO design and delivery

All ASO molecules used were synthesized (100nmol scale) at Integrated DNA Technologies (IDT) with a 2’ O-methyl modification (Table 2). ASOs were added at a final concentration of 10uM, which was introduced with the transfection reagent TransIT-TKO (Mirius, MIR2250). For example, TransIT-TKO was added to Opti-MEM (ThermoFisher Scientific, 31985062) and mixed gently by pipetting up and down. For every 100uL of Opti-MEM-ASO mix, 3uL of TransIT-TKO was used. The ASOs were then added to the Opti-MEM-TransIT-TKO mix at the desired concentration and mixed gently by pipetting and incubated at room temperature for 45 minutes. After 45 minutes, the ASO transfection mix was added dropwise and gently shaken to evenly distribute the ASO.

ASOs were added either to iPSCs or cardiac organoids. For iPSCs, ASOs were added and 48 hours later media was changed to mTeSR1. For cardiac organoids, ASOs were added 14-22 days after the start of differentiation and media was changed 48 hours later. The sequences of all ASOs used in this study are shown in Table S2.

#### iPSC-Cardiac organoid differentiation

To differentiate iPSCs into cardiac organoids cells were dissociated using Accutase (ThermoFisher Scientific, A1110501) for EB formation. After dissociation, cells were centrifuged at 300xg for 3 min and resuspended in mTeSR1 medium (StemCell Tech, 85850) containing 10uM ROCK inhibitor (Y-27632 dihydrochloride, Tocris, 1254). iPSCs were counted and seeded at 5,000 cells/well in a 96-well ultra-low binding, U-shaped-bottom microplate (Corning, 4515) or 5,000 cells per microwell when using the AggreWell 800 plates (StemCell Tech, 34825). The plate was centrifuged at 100xg for 3 min and placed in an incubator at 37C, 5% CO_2_. After 3-5 days EBs were ready for cardiac organoid differentiation. EBs will have round and smooth edges when they are ready for differentiation. Cardiac differentiation was started by the addition of Cardiac differentiation media (RPMI 1640/B-27) (CDM) (Fisher, 11875119, A1895601), minus insulin containing CHIR99021 (StemCell Tech, 72054) at a final concentration of 6uM. After 2 days media was changed to CDM, minus insulin containing IWP2 (Tocris, 3533) at a final concentration of 5uM for 2 days. After 2 days media was changed to CDM, minus insulin for an additional 2 days followed by the addition of maintenance media (RPMI 1640/B27, with insulin) (Fisher, 17504044). Media was changed every 2-4 days with maintenance media for the duration of the experiment.

#### Immunofluorescence

Cardiac organoids were fixed in 4% PFA solution (Sigma, 252549) overnight at 4C. Fixation was followed by washes in 1X PBS (Sigma, D8537) and permeabilization using 1X PBS with 0.2% TX-100 (Sigma, X100) for 30 min at room temperature. After permeabilization, blocking was performed overnight at 4C in blocking solution (1X PBS, 5% FBS (ThermoFisher Scientific, 10082147), 0.2% TX-100, 2.5% BSA (Sigma, A9418)). The next day cardiac organoids were washed 3×3 min in 1X PBS containing 0.1% BSA followed by the addition of primary antibody solution for 2-3 days at 4C. Primary antibody exposure was followed by 5×3 min washes and incubation with secondary antibodies for 24 hours at 4C in the dark. After secondary antibody incubation, cardiac organoids were washed 5×3 min in 1X PBS and imaged using a fluorescence microscope (Keyence BZ-X810). Antibody solution was 1:10 of blocking solution in 1X PBS. Primary antibody (Dystrophin, Abcam, ab15277) (Cardiac Troponin T Monoclonal Antibody, ThermoFisher Scientific, MA5-12960) was used at 1:100. Secondary antibody (Goat anti-Rabbit Alexa Fluor 488, Goat anti-Mouse Alexa Fluor 594) (Fisher, A11034, A11032) was used at 1:400. Immunostaining was as described above for iPSCs.

#### Calcium imaging

Cardiac organoids were loaded with 10uM Fluo-4 AM (ThermoFisher Scientific, F14217) with 1.25mM probenecid (Fisher, P36400) and 0.02% pluronic F-127 (Fisher, P6867) added directly to the maintenance media (RPMI 1640/B27, with insulin) for 45 min at 37C. The cardiac organoids were washed, and maintenance media added back and incubated at 37C for an additional 30 min before responses were measured using a fluorescence microscope (Keyence BZ-X810) using the FITC filter set. Quantification of cardiac contraction was performed using Musclemotion, a free open-source software for ImageJ.

#### Ethics statement

The Institutional Review Board (IRB) of Children’s Mercy Research Institute gave ethical approval for this work (Studies #00002465 and #11120514). All methods were carried out in accordance with relevant guidelines and regulations.

## Acknowledgements

This study was supported by generous philanthropic contributions to the Children’s Mercy Research Institute and the Genomic Answers for Kids program.

## Authors’ contributions

STY and JCM conceived the study. JCM designed, performed, and analyzed experiments. DAL manages the GA4K biobank. TP and EGF assisted in patient selection. STY and JCM wrote the manuscript. STY supervised the study. All authors read and approved the final manuscript.

## Supplemental Figure Legends

**Supplemental Figure 1. Generating iPSC models from patient fibroblasts. a)** Patient fibroblasts. **b)** An iPSC colony derived from patient fibroblasts shown in Figure 1a.

**Supplemental Figure 2. Cardiac organoids derived from DMD patient 1**. Cardiac troponin T expression in cardiac organoids derived from patient 1 and treated with ASOs matching the sequence of existing ASO therapeutics.

**Supplemental Figure 3. Arrhythmic contraction in cardiac organoids derived from patient 1**. Contraction of individual DMD patient-derived cardiac organoids (patient 1), rescaled data from Figure 3 to highlight arrhythmic and low intensity contractions in untreated organoids and organoids treated with a nontargeting ASO.

**Supplemental Figure 4. Sustained dystrophin expression in ASO-treated cardiac organoids derived from DMD patient 1**. Dystrophin expression in cardiac organoids derived from patient 1 and treated with ASOs matching the sequence of existing ASO therapeutics.

**Supplemental Figure 5. Cardiac organoids derived from DMD patients 2a/2b. a)** Cardiac troponin T expression in cardiac organoids derived from patient 2a and treated with patient-specific ASOs. **b)** Cardiac troponin T expression in cardiac organoids derived from patient 2b and treated with patient-specific ASOs.

**Supplemental Figure 6. Arrhythmic contraction in cardiac organoids derived from patients 2a/b. a)** Contraction of individual DMD patient-derived cardiac organoids (patient 2a), rescaled data from Figure 4 to highlight arrhythmic and low intensity contractions in untreated organoids and organoids treated with a nontargeting ASO. **b)** Contraction of individual DMD patient-derived cardiac organoids (patient 2b), rescaled data from Figure S8 to highlight arrhythmic and low intensity contractions in untreated organoids and organoids treated with a nontargeting ASO.

**Supplemental Figure 7. Sustained dystrophin expression in ASO-treated cardiac organoids derived from DMD patients 2a/2b. a)** Dystrophin expression in cardiac organoids derived from patient 2a and treated with patient-specific ASOs. **b)** Dystrophin expression in cardiac organoids derived from patient 2b and treated with patient-specific ASOs.

**Supplemental Figure 8. Preclinical evaluation of patient-specific ASOs in additional patient-derived organoids. a)** Dystrophin expression in DMD patient-derived cardiac organoids (patient 2b) treated with patient-specific ASOs. **b)** Contraction of individual DMD patient-derived cardiac organoids (patient 2b) treated with patient-specific ASOs. **c)** Time course analysis of restored contraction in DMD patient-derived cardiac organoids (patient 2b) treated with patient-specific ASOs.

**Supplemental Table 1. Summary of iPSC lines**.

**Supplemental Table 2. ASO sequences and targets**. All ASOs in this study were synthesized as fully 2’-O-methyl-modified RNAs.

**Supplemental Video 1. Fluo-4 imaging of a cardiac organoid**. Cardiac organoid generated from a commercially available iPSC line treated with a nontargeting ASO prior to differentiation.

**Supplemental Video 2. Fluo-4 imaging of a cardiac organoid**. Cardiac organoid generated from a commercially available iPSC line treated with an ASO targeting the cardiac troponin T AUG translation start site prior to differentiation.

**Supplemental Video 3. Fluo-4 imaging of a cardiac organoid**. Cardiac organoid generated from a commercially available iPSC line treated with an ASO targeting a splice donor site in the cardiac troponin T gene prior to differentiation.

**Supplemental Video 4. Fluo-4 imaging of a cardiac organoid**. Cardiac organoid generated from a commercially available iPSC line treated with an ASO targeting a splice acceptor site in the cardiac troponin T gene prior to differentiation.

**Supplemental Video 5. Fluo-4 imaging of a cardiac organoid**. Cardiac organoid generated from a commercially available iPSC line.

**Supplemental Video 6. Fluo-4 imaging of a cardiac organoid**. Cardiac organoid generated from an iPSC line derived from Patient 1.

**Supplemental Video 7. Fluo-4 imaging of a cardiac organoid**. Cardiac organoid generated from an iPSC line derived from Patient 1 and treated with a nontargeting ASO.

**Supplemental Video 8. Fluo-4 imaging of a cardiac organoid**. Cardiac organoid generated from an iPSC line derived from Patient 1 and treated with a DMD exon 45 skipping ASO matching the sequence of an FDA-approved ASO.

**Supplemental Video 9. Fluo-4 imaging of a cardiac organoid**. Cardiac organoid generated from an iPSC line derived from Patient 1 and treated with a DMD exon 45 skipping ASO matching the sequence of an ASO in phase II clinical trials.

**Supplemental Video 10. Fluo-4 imaging of a cardiac organoid**. Cardiac organoid generated from an iPSC line derived from Patient 2a.

**Supplemental Video 11. Fluo-4 imaging of a cardiac organoid**. Cardiac organoid generated from an iPSC line derived from Patient 2a and treated with a nontargeting ASO.

**Supplemental Video 12. Fluo-4 imaging of a cardiac organoid**. Cardiac organoid generated from an iPSC line derived from Patient 2a and treated with ASO-1 targeting a pathogenic intronic variant in the DMD gene.

**Supplemental Video 13. Fluo-4 imaging of a cardiac organoid**. Cardiac organoid generated from an iPSC line derived from Patient 2a and treated with ASO-2 targeting a pathogenic intronic variant in the DMD gene.

**Supplemental Video 14. Fluo-4 imaging of a cardiac organoid**. Cardiac organoid generated from an iPSC line derived from Patient 2b.

**Supplemental Video 15. Fluo-4 imaging of a cardiac organoid**. Cardiac organoid generated from an iPSC line derived from Patient 2b and treated with a nontargeting ASO.

**Supplemental Video 16. Fluo-4 imaging of a cardiac organoid**. Cardiac organoid generated from an iPSC line derived from Patient 2b and treated with ASO-1 targeting a pathogenic intronic variant in the DMD gene.

**Supplemental Video 17. Fluo-4 imaging of a cardiac organoid**. Cardiac organoid generated from an iPSC line derived from Patient 2b and treated with ASO-2 targeting a pathogenic intronic variant in the DMD gene.

## References

1. Thakur, S., Sinhari, A., Jain, P. & Jadhav, H. R. A perspective on oligonucleotide therapy: Approaches to patient customization. Front Pharmacol 13, 1006304 (2022).

2. Kim, J. et al. Patient-Customized Oligonucleotide Therapy for a Rare Genetic Disease. New Engl J Med 381, 1644–1652 (2019).

3. Doss, M. X. & Sachinidis, A. Current Challenges of iPSC-Based Disease Modeling and Therapeutic Implications. Cells 8, 403 (2019).

4. Wu, J. C. et al. Towards Precision Medicine With Human iPSCs for Cardiac Channelopathies. Circ Res 125, 653–658 (2019).

5. Cohen, A. S. A. et al. Genomic answers for children: Dynamic analyses of >1000 pediatric rare disease genomes. Genet Med 24, 1336–1348 (2022).

6. Ye, H. & Wang, Q. Efficient Generation of Non-Integration and Feeder-Free Induced Pluripotent Stem Cells from Human Peripheral Blood Cells by Sendai Virus. Cell Physiol Biochem 50, 1318–1331 (2018).

7. Bang, J. S. et al. Optimization of episomal reprogramming for generation of human induced pluripotent stem cells from fibroblasts. Anim Cells Syst 22, 132–139 (2018).

8. Nishii, K. et al. Targeted disruption of the cardiac troponin T gene causes sarcomere disassembly and defects in heartbeat within the early mouse embryo. Dev Biol 322, 65–73 (2008).

9. Umemoto, N. et al. Fluorescent-Based Methods for Gene Knockdown and Functional Cardiac Imaging in Zebrafish. Mol Biotechnol 55, 131–142 (2013).

10. Dai, Y. et al. Troponin destabilization impairs sarcomere-cytoskeleton interactions in iPSC-derived cardiomyocytes from dilated cardiomyopathy patients. Sci Rep-uk 10, 209 (2020).

11. Syed, Y. Y. Eteplirsen: First Global Approval. Drugs 76, 1699–1704 (2016).

12. Heo, Y.-A. Golodirsen: First Approval. Drugs 80, 329–333 (2020).

13. Dhillon, S. Viltolarsen: First Approval. Drugs 80, 1027–1031 (2020).

14. Shirley, M. Casimersen: First Approval. Drugs 81, 875–879 (2021).

15. Hoffman, E. P. et al. Novel Approaches to Corticosteroid Treatment in Duchenne Muscular Dystrophy. Phys Med Rehabil Cli 23, 821–828 (2012).

16. Mercuri, E., Bönnemann, C. G. & Muntoni, F. Muscular dystrophies. Lancet 394, 2025–2038 (2019).

17. Ito, K. et al. Renadirsen, a Novel 2′OMeRNA/ENA® Chimera Antisense Oligonucleotide, Induces Robust Exon 45 Skipping for Dystrophin In Vivo. Curr Issues Mol Biol 43, 1267–1281 (2021).

18. Waldrop, M. A. et al. Intron mutations and early transcription termination in Duchenne and Becker muscular dystrophy. Hum. Mutat. 43, 511–528 (2022).

19. Putten, M. et al. Low dystrophin levels increase survival and improve muscle pathology and function in dystrophin/utrophin doubleLknockout mice. Faseb J 27, 2484–2495 (2013).

20. Wells, D. J. What is the level of dystrophin expression required for effective therapy of Duchenne muscular dystrophy? J Muscle Res Cell M 40, 141–150 (2019).

21. Wang, Y.-X. & Deng, M. Medical imaging in new drug clinical development. J Thorac Dis 2, 245–52 (2011).

22. Réda, C., Kaufmann, E. & Delahaye-Duriez, A. Machine learning applications in drug development. Comput Struct Biotechnology J 18, 241–252 (2020).

23. Wright, C. F., FitzPatrick, D. R. & Firth, H. V. Paediatric genomics: diagnosing rare disease in children. Nat Rev Genet 19, 253–268 (2018).

