## Supplementary figures and images for "Rapid and scalable preclinical evaluation of personalized antisense oligonucleotide therapeutics using organoids derived from rare disease patients"

### Figure S1

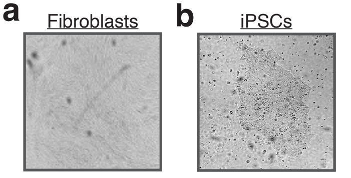

### Figure S2

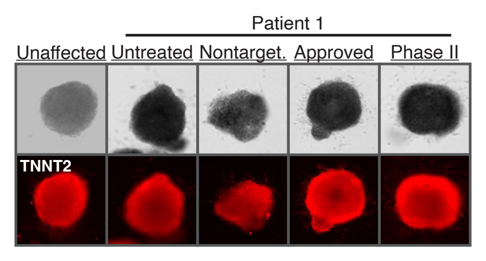

### Figure S3

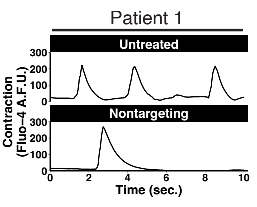

### Figure S4

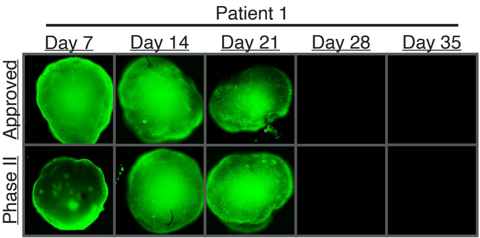

### Figure S5

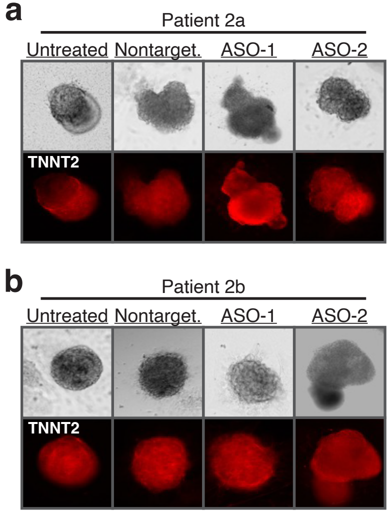

### Figure S6

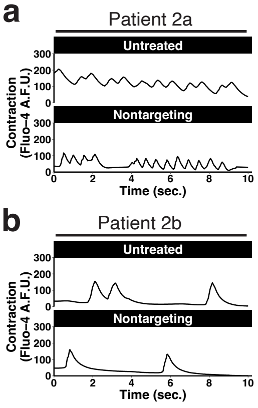

### Figure S7

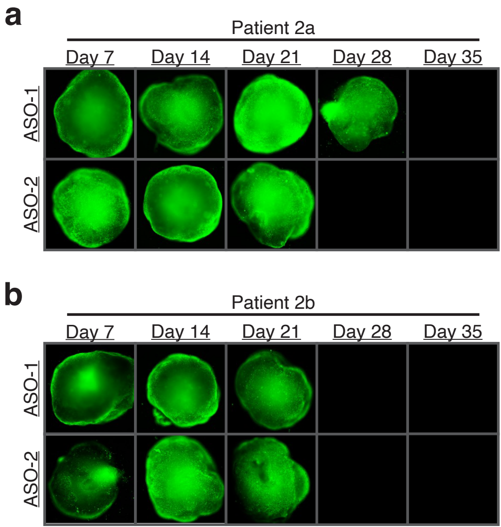

### Figure S8

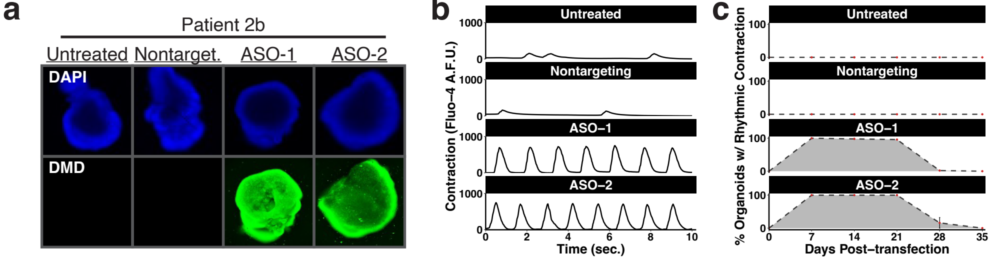
